# Persistent Homology in Medical Image Processing: A Literature Review

**DOI:** 10.1101/2025.02.21.25322669

**Authors:** Daniel Brito-Pacheco, Panos Giannopoulos, Constantino Carlos Reyes-Aldasoro

## Abstract

Medical image analysis has experienced significant advances with the integration of machine learning, deep learning, and other mathematical and computational methodologies into the pipelines of data analysis. One methodology that has received less attention is Persistent Homology (PH), which comes from the growing field of Topological Data Analysis and has the ability to extract features from data at different scales and build multi-scale summaries. In this work, we present a systematic review of PH applied in medical images. To illustrate the potential of PH, we introduce the main concepts of PH and demonstrate with an example of histopathology. Fifteen articles where PH was applied to medical image analysis tasks such as segmentation and classification were selected and reviewed. It was observed that PH is very versatile, as it can be applied in many different contexts and to different data types, whilst also showing great potential in increasing model accuracy in both classification and segmentation. It was also observed that image segmentation predominantly uses basic level-set filtration to calculate PH, while classification takes various approaches using filtration on more complex structures built from data. This review highlights PH as an important tool that can further advance medical image analysis.

## 1 Introduction

Medical image processing has seen steady growth since the 1970s when algorithmic approaches applied to analyse medical images started to show promising results [1–3]. Today, thousands of algorithms used to analyse medical images are published every year to much acclaim in areas such as histology [4] and radiology [5, 6]. Many of the algorithms described in these papers exploit techniques like Machine Learning (ML) [7], Mathematical Morphology [8], Fourier Analysis [9], and, more recently, Deep Learning [10–19], which is dominating the literature [20], for a variety of tasks such as nuclear detection, segmentation, classification and counting [21], predicting survival in cancer [22], out-of-distribution detection and localization [23].

A series of mathematical techniques that have received less attention are those related to Persistent Homology (PH) [24–26]. PH considers data as a multi-scale representation of topological features and enables the analysis of higher dimensional representations where the data may have a predetermined *shape* [27]. PH allows for the identification of this shape from the data and has already been successfully used in areas as diverse as quantification of airway structures [28], analysis of functional brain activity [29], analysis of glass structure [30], the study of mitochondrial network connectivity [31], the segmentation of cell mitochondria [32], and the detection and segmentation of cell nuclei [33].

As such, PH has the potential to be a powerful instrument in the study of the immune system, especially when combined with other tools into image analysis algorithms and pipelines and may be particularly useful with higher dimensional data. In this review, a comparison and examination of the existing literature on PH in medical image processing is performed the use of this technique in the analysis of medical dataset is explored.

## 2 Background

### 2.1 Persistent Homology

The Merriam-Webster dictionary presents four definitions of Homology: (1) similarity due to a common origin, (2) similarity of traits reflecting common descent and ancestry, (3) similarity of nucleotide or amino acid sequence, and (4) a branch of the theory of topology concerned with partitioning space into geometric components. Persistent Homology is closest to the fourth definition, which is the focus of this work, but the first two definitions are also relevant, as homology implies similarity of mathematical properties. It is interesting to notice that outside the topological context, there is discrepancy in the understanding of homology. Marabotti and Facchiano [34] insist that homology should only refer to *having a common evolutionary origin* and not as similarity that can be quantified, but only around 57% of articles published in 2007 used this and the other 43% used homology as a measure of similarity.

Persistent Homology is one of the main tools in the mathematical field of Topological Data Analysis (TDA), a field which uses concepts from the mathematical branch of Topology to create statistical models for analyzing data [35–37]. The essence of topology is to consider only some aspects of objects and completely disregard others. This was the approach taken by Euler to solve the problem of the seven bridges of Königsberg [38], which required to walk over each of the seven bridges of the city of Königsberg in Prussia exactly once and returning to the starting point. To solve this problem, the length, width or material of the bridges are irrelevant. The only important information is which islands of the city are connected by which bridges. The property of connectivity does not change if the bridges are deformed or change shape as long as they connect the same points in a map. Following this logic, a sphere can be deformed and maintain the same topological characteristics as an ellipsoid, a doughnut can be deformed and maintain the same characteristics as a cup with one handle or indeed a section of a blood vessel or the intestines. The key difference between the sphere and the doughnut relies in the number of *holes* each object has. A cup with two handles is different to the doughnut as it contains two holes, but it is similar or homologous to a double torus or a cross section of a heart as illustrated in Fig. 1.

**Fig. 1.**
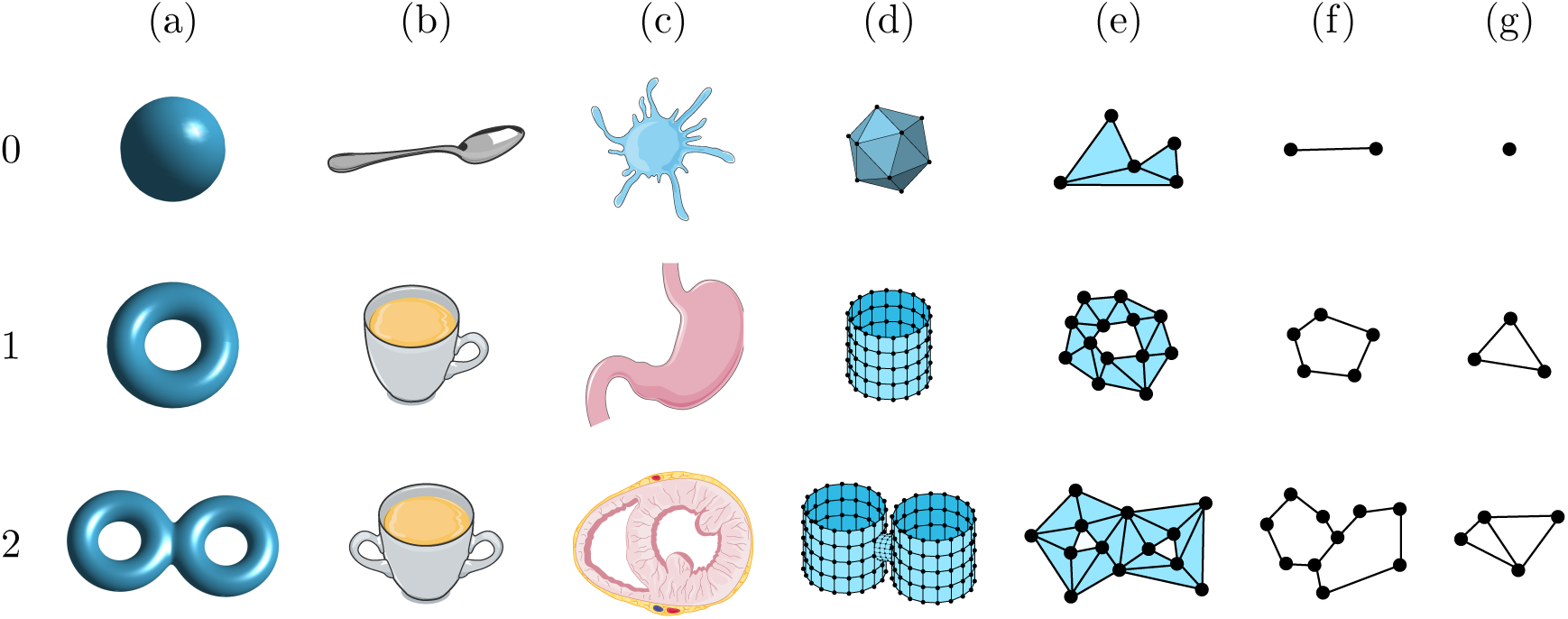
Different objects grouped by the number of *holes* that they have. Row 0, no holes: (a) sphere, (b) spoon, (c) cartoon of a dendritic cell, (d) dodecahedron, (e) a union of triangles with no holes, (f) two points joined by an edge, (g) single point. Row 1, one hole: (a) solid torus, (b) cup with a single handle, (c) cartoon of a stomach, (d) cylinder, (e) union of triangles surrounding a region of 2D space, (f) points joined by a cycle of 5 edges, (g) triangle not filled in. Row 2, two holes: (a) two-holed torus, (b) cup with two handles, (c) cartoon of a heart cross-section, (d) two cylinders joined together, (e) union of triangles surrounding two different regions, (f) points joined by two cycles of edges, (g) two triangles not filled in, joined by an edge.

### 2.2 Simplicial Complexes and persistence

A *simplicial complex* is a mathematical structure that can be used to represent complex objects as topological spaces in a simplified manner. The complex is formed of points, edges, triangles, tetrahedra, etc., all of which are collectively known as *simplices*. Persistent Homology tracks the topological changes that occur to a topological space as it deforms according to a parameter, for instance, by slicing a 3D object *e.g.*, a plant (Fig. 2 (a)), or a bronchial tree (Fig. 2 (b)) at different levels. The parameter would be the height where the object is sliced and the deformation would follow to what is being sliced: roots, stem, branches and leaves, etc. (or larynx, trachea, bronchi, etc.). In the case of a greyscale image, an intensity threshold on the brightness of pixels could be lowered or raised progressively, creating a filtration from the pixels that are brighter than the threshold as illustrated by Fig. 2 and Fig. 3. As the threshold moves, regions above the threshold (Fig. 3(a)) become the components (Fig. 3(b)) and holes (Fig. 3(c)), which appear, or are *born* and represented by a bar in the Figure. The simplification of the components becomes the simplicial complex (Fig. 3(d)). When two components merge into a single component, one of them disappears, or *dies*. Typically, the oldest component survives (the one that was born earlier in the filtration). When a hole is filled in, it disappears and is also said to die at that time point. The birth and death of components and holes is encoded in a scatter plot known as the *Persistence Diagram* (Fig. 3(e)). The horizontal coordinates of circles (components) or triangles (holes) correspond to the time point of birth, and the vertical coordinate to the time point of death. The case may arise where two components (or holes) are born and die at the same times, thus leading to two different points having the same coordinates in the persistence diagram; there exist other visualisations, such as persistence barcodes, that do not have this problem [39]. The persistence diagram can be understood as a summary of the topological changes that occur through the sequence of deformations. It is a topological signature and can be used for comparing two shapes using well-known distance metrics [40, 41]. Finally, the number of components and holes in a complex are summarised metrics called *Betti Numbers*. The 0-th Betti Number (*β*_0_) of a simplicial complex counts the amount of components, while the 1-st Betti Number (*β*_1_) counts the amount of holes.

**Fig. 2.**
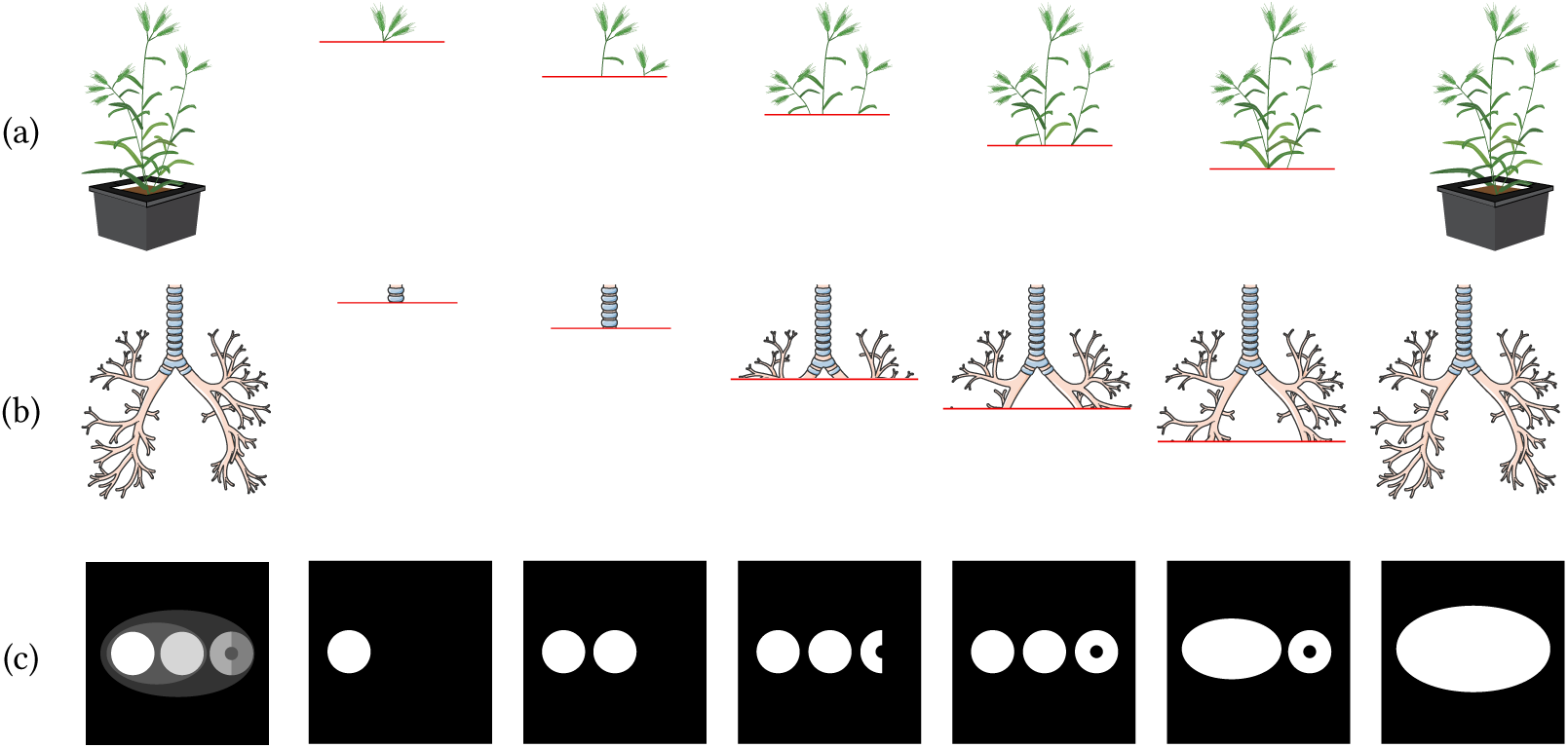
Illustration of filtration process by thresholding. (a) A height threshold is applied to a plant. At each position of the threshold, the plant would be sliced and the filtered data would correspond to the parts of the plant visible on that plane. (b) Similarly, a height threshold is applied to a bronchial tree, thus letting different parts of the bronchial tree visible at each height. (c) A pixel intensity threshold is applied to a greyscale image, generating six binary images. In this case, pixels above a certain threshold are coloured white, while pixels below the threshold are coloured in black.

**Fig. 3.**
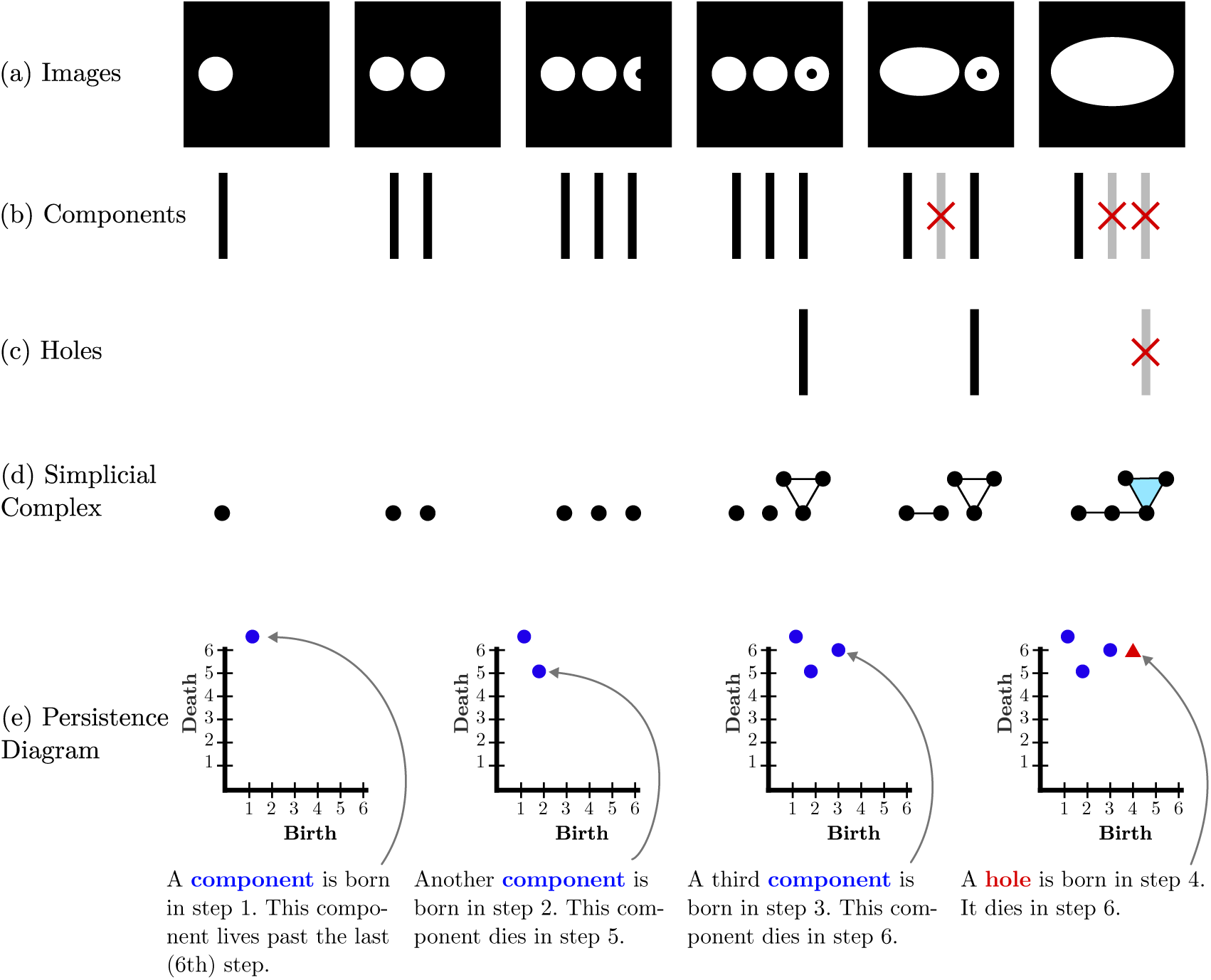
Illustration of the main concepts of Persistence Homology. In this example, the birth and death of the holes and components are tracked within the six binary images created from the intensity threshold in Fig 2 (c) and noted in the persistence diagrams. (a) Six binary images with pixels that are above a threshold that is progressively decreased, from Fig 2 (c). (b) A black bar is added to illustrate the birth of a component and when the component dies, it is marked with a red cross and changed to grey. (c) The birth and death of holes is marked in the same way as for components. (d) Representation of the components as a simplicial complex. A point is added for each connected component, except if there is a hole, in which case a cycle of 3 points and 3 edges is added; the cycle is filled in when the hole dies. (e) Step by step formation of the persistence diagram. The first component that appears at step 1 will continue to live beyond the last step (6). The second component is born in step 2 and dies in step 5 when it merges with the first component. The oldest component always survives a merger. The third component appears in step 3 and dies in step 6. A hole is born in step 4 and dies in step 6.

It is important to note that these definitions are informal and without mathematical notation. For a more formal introduction, the reader is referred to [24].

### 2.3 Level-Set Filtration

One of the most common techniques to calculate PH is through *level-set filtration*, which was briefly described in subsection 2.2. Consider a greyscale image of dimensions 8 × 8 where each pixel can have values between 0 and 255 (Fig. 4(a)). An intensity level threshold is used to segment the image into regions, starting at 255 and decreasing to 0 and as the threshold is lowered (Fig. 4(c)), new regions are born and then die as they merge (Fig. 4(d)). In a merger, the older component absorbs the one that dies. Next, the simplicial complexes are overlaid on the binary images (Fig. 4(e)). A vertex (0-simplex) is placed at each white pixel, neighbouring white pixels (including diagonal neighbours) are joined through an edge (1-simplex), and a triangle (2-simplex) is added when three edges create a cycle, which can be filled as in the first column of (Fig. 4(e)). Finally, the binary regions are removed to show only the simplicial complex of each step (Fig. 4(f)).

**Fig. 4.**
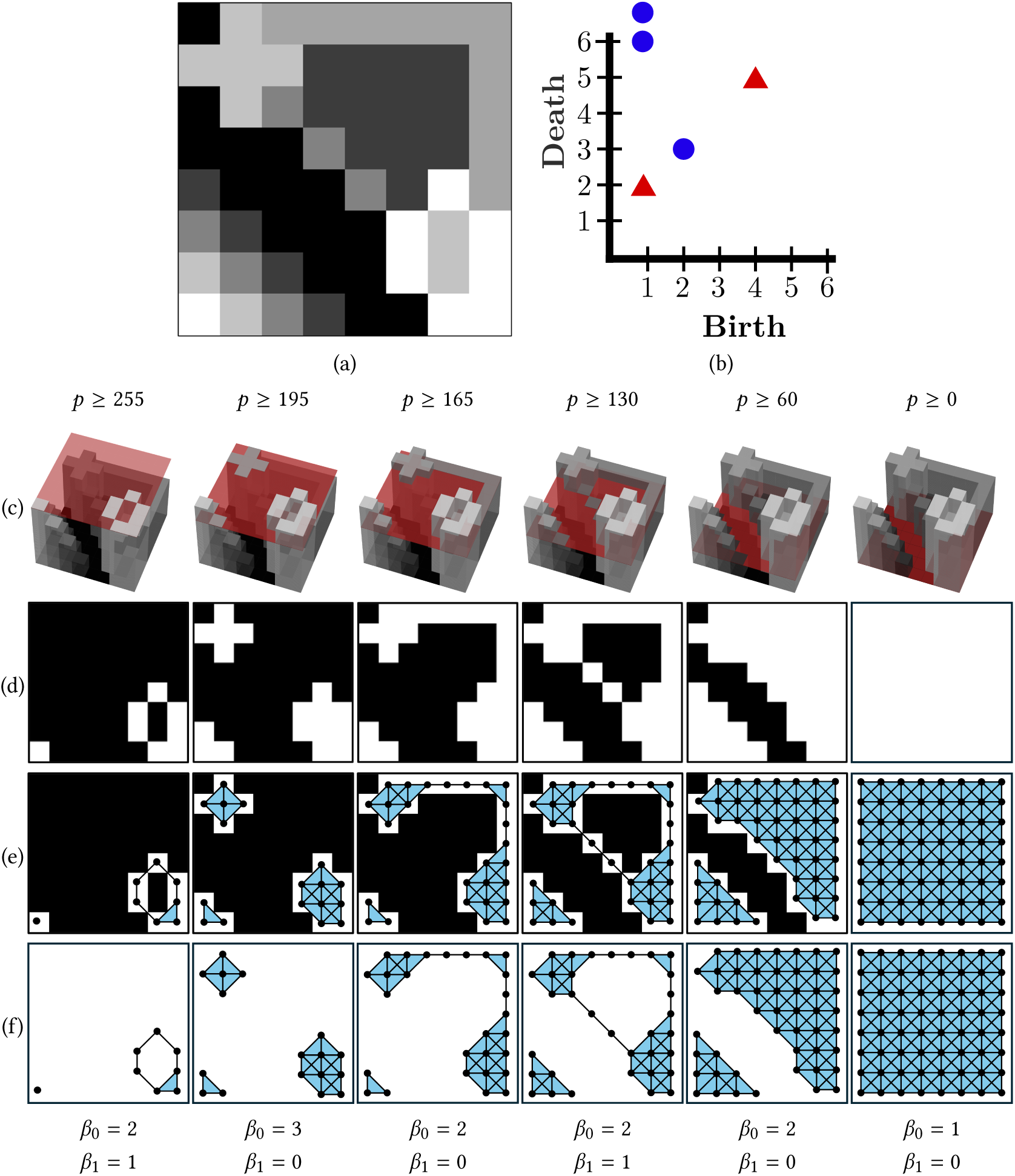
Illustration of the filtration and PH calculation of a greyscale image with level-set thresholds. (a) Original greyscale image with pixel intensities in the range [0, 255]. (b) Persistence diagram. (c) 3D representation of the greyscale image with thresholds illustrated by a red transparent surface. (d) Binary images of pixels above the threshold. (e) Filtration overlaid on the binary images. A vertex (point) is placed at each white pixel. An edge is added between neighbouring pixels. A triangle is added when cycles of three edges are formed. (f) Filtration with the pixels removed. Betti numbers for each case are added at the bottom of the figure.

When the filtration process is finished, births and deaths are encoded into a Persistence Diagram (Fig. 4(b)). In this diagram, each point corresponds to a topological feature. The horizontal coordinate of the point corresponds to its time of birth while the vertical coordinate encodes the time of death.

It is important to highlight that there are other filtration strategies (like slicing a plant), however, level-set filtration is probably the most popular [42–44]. In fact, the way in which the filtration is built is where most applications of PH differ from one another and will be considered in the review below.

### 2.4 Persistent Homology in Histology: An Example

In this section, PH is applied to histology Whole Slide Images (WSI) of colorectal cancer (CRC) patients to illustrate how it is able to capture differences in textures and patterns of tissues. This example serves also as a way to show how PH can be directly useful in extracting features from medical images.

The dataset used for this example is NCT-CRC-HE-100K [45]. The set of images contains around 100,000 patches from H&E-stained human cancer tissue WSIs. All images in the set are 224 × 224 pixels, at a resolution of 5*μm* per pixel and normalized with the Macenko method [46]. Each patch belongs to one of nine possible tissue classes: adipose tissue (ADI), background (BACK), debris (DEB), lymphocytes (LYM), mucus (MUC), smooth muscle (MUS), normal colon mucosa (NORM), cancer-associated stroma (STR), and CRC epithelium (TUM). Fig. 5 shows five example images from each tissue class in the dataset.

**Fig. 5.**
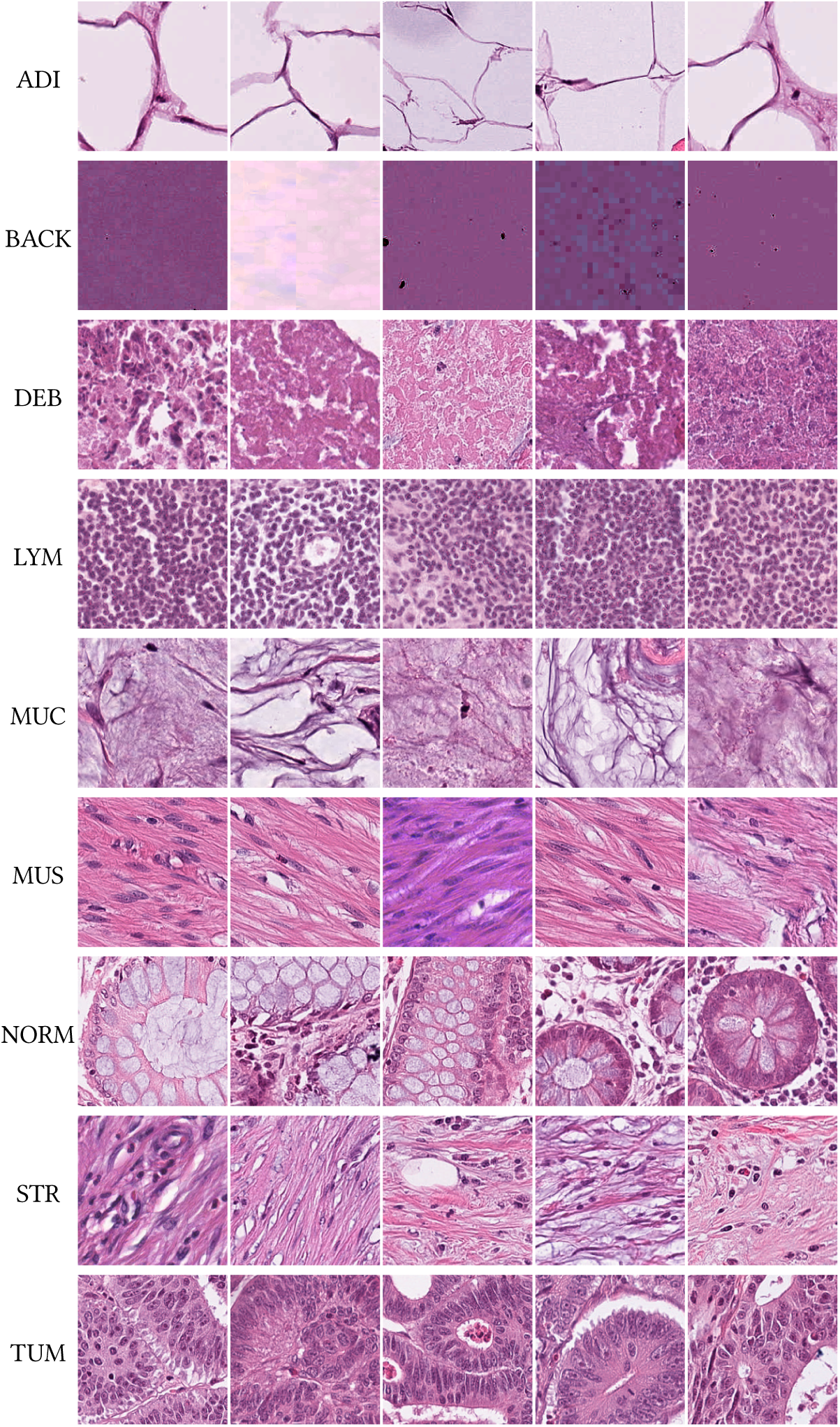
Illustration of histological tissues of 9 classes. Each row contains five representative examples from each of the nine classes in the NCT-CRC-HE-100K dataset [45]. ADI: adipose tissue; BACK: background; CRC: colorectal cancer; DEB: debris; LYM: lymphocytes; MUC: mucus; MUS: smooth muscle; NORM: normal colon mucosa; STR: cancer-associated stroma; TUM: colorectal adenocarcinoma epithelium.

Nine persistence diagrams, one for each type of tissue, are illustrated in Figure 6. Each patch is converted to greyscale and inverted (dark regions are made bright). Then, a 5 × 5 median filter is applied to the resulting image to reduce noise. Finally, the persistence diagram is computed by using the level-set filtration introduced in subsection 2.3, i.e., by applying a threshold that sweeps all the intensities of the greyscale patch to track the birth and death of components and holes. It is important to notice how the persistence diagrams capture the differences in textures and patterns visible in the different patches, in particular:

- Adipose tissue (ADI) forms bright bubbles (in light pink) with dark borders (in purple). In the greyscale inverted image, this is seen as dark regions surrounded by a light-grey / white membrane. This description is captured by the large group of blue circles concentrated towards the bottom of the persistence diagram: as the intensity threshold drops from 255, the components coalesce into a single component which persists for a long time (a blue circle with high persistence).
- The number of holes depends on the dark areas surrounded by brighter pixels: the amount of red triangles in the persistence diagram computed from the LYM patch is clearly greater than those calculated from the ADI patch.
- The greyscale version of the TUM patch has a few very dark spots (white in the colour version). This is captured by the persistence of red triangles (how high they are compared to the diagonal).
- The BACK patch has very little variation in pixel intensities, which is clearly identified in the tight clusters that both components and holes form in the persistence diagram.

**Fig. 6.**
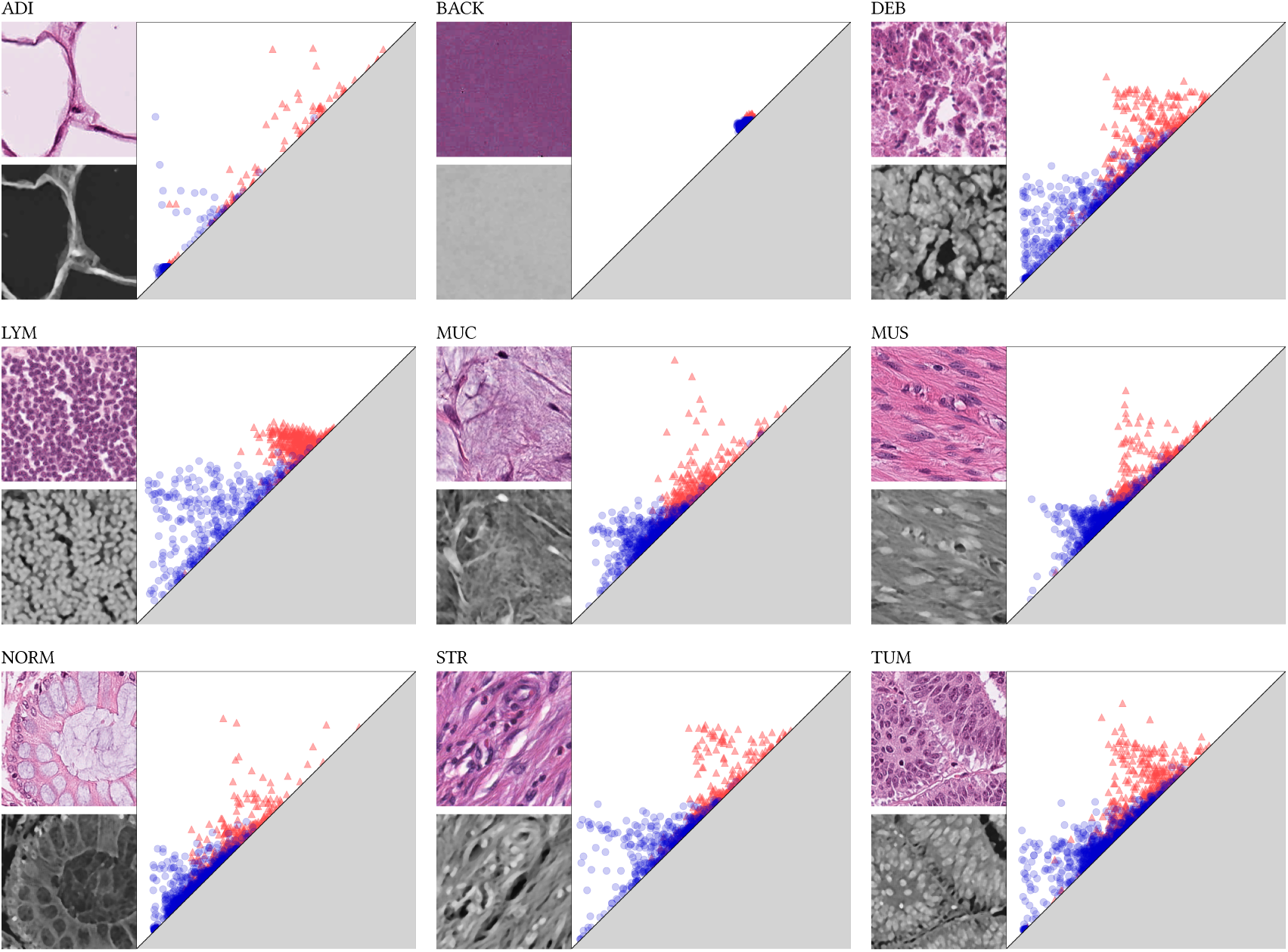
Illustration of the persistence diagrams of the histological tissues presented in Figure 5. One representative patch from each of the classes (ADI, BACK, DEB, LYM, MUC, MUS, NORM, STR, TUM) from the NCT-CRC-HE-100K dataset [45] is converted to grayscale and inverted. Noise is removed on the grayscale image by applying a 5 × 5 median filter. A persistence diagram is calculated on the smoothed grayscale image where blue circles are components and red triangles are holes. The distribution of the scatterplots in the persistence diagram capture differences in the textures of different tissues; for instance, the persistence diagram of adipose tissue sees most components born and die early since bright areas in the grayscale join relatively quickly as the threshold lowers; this contrasts with Lymphocytes where the components take a longer time to join into one single component. ADI: adipose tissue; BACK: background; CRC: colorectal cancer; DEB: debris; LYM: lymphocytes; MUC: mucus; MUS: smooth muscle; NORM: normal colon mucosa; STR: cancer-associated stroma; TUM: colorectal adenocarcinoma epithelium.

The previous descriptions refer to characteristics of single patches. However, statistical features, such as the mean persistence of the components, can be calculated from each persistence diagram. In Figure 7, four statistical features are calculated from a sample of 500 patches of each class and plotted to illustrate clusters emerging from the topological features. The features that are calculated are: total number of components, mean persistence of components, standard deviation of the persistence of holes, median death of holes. Figure 7 (a) shows a scatterplot of the first two, while Figure 7 (b) shows a scatterplot of the third and last features. A representative patch from seven of the nine classes is shown connected to the point in the scatterplot corresponding to the features calculated from it. In this figure, it is noticeable that the cluster for the LYM class is very distinctive on both plots. This is likely due to the characteristic size and distribution of lymphocytes, which, in patches of consistent size, result in a relatively uniform tissue texture. Additionally, in Figure 7 (a) two distinct groups of the ADI class are distinguishable. The representative patches show that this may be due to two different subclasses of adipose tissue, one that forms with a darker histogram than the other.

**Fig. 7.**
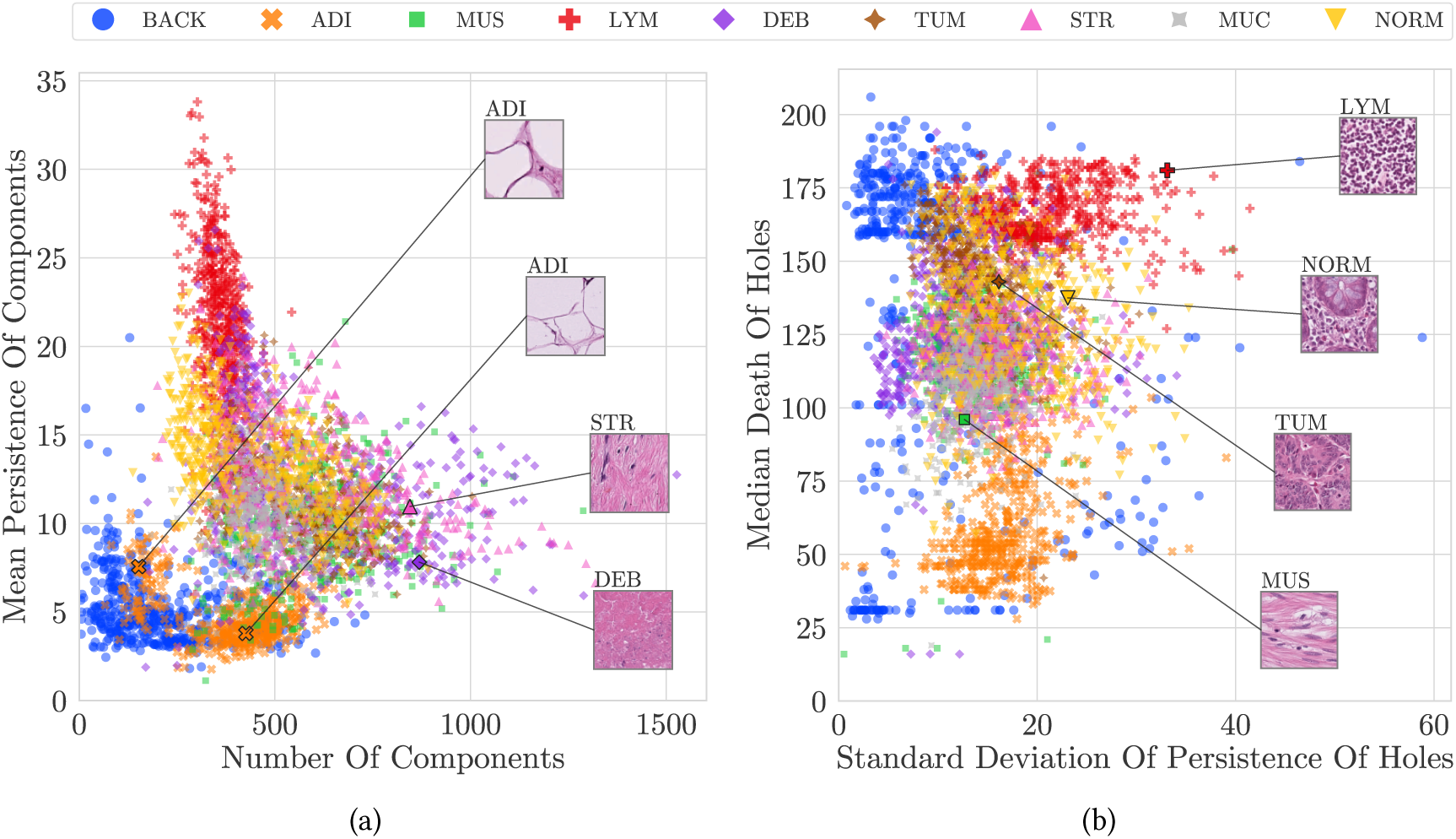
Illustration of metrics extracted from the persistence diagrams. Different statistical attributes were calculated from the persistence diagrams of 500 representative patches of each tissue. Namely, the total number of components, mean persistence of components, standard deviation of persistence of holes and median death of holes were calculated. (a) Number of components against mean persistence of components from each of the nine classes of tissue. (b) Standard deviation of the persistence of holes against median death of holes. In both cases it is possible to observe how PH features such as these could be used to distinguish between tissue types, as, for example, LYM patches (red crosses) show a tight cluster easily distinguishable in both plots. The background (blue circles) is concentrated in the lower left in (a), but spread over the vertical axis in (b), this may be due to variations on the intensity (dark/bright) that are visible in Figure 5. It can also be observed that ADI seems to be split in two regions in (a). ADI: adipose tissue; BACK: background; DEB: debris; LYM: lymphocytes; MUC: mucus; MUS: smooth muscle; NORM: normal colon mucosa; STR: cancer-associated stroma; TUM: colorectal adenocarcinoma epithelium.

## 3 Materials and Methods

A systematic review to identify the use of PH in Medical Image Processing literature was performed following the Preferred Reporting Items for Systematic Reviews and Meta-Analyses (PRISMA) [47, 48] and the guidelines proposed in [49]. The steps of the review are illustrated in Fig. 8. The only database considered for the review was Medline, which was queried through the search engine PubMed (https://pubmed.ncbi.nlm.nih.gov/).

**Fig. 8.**
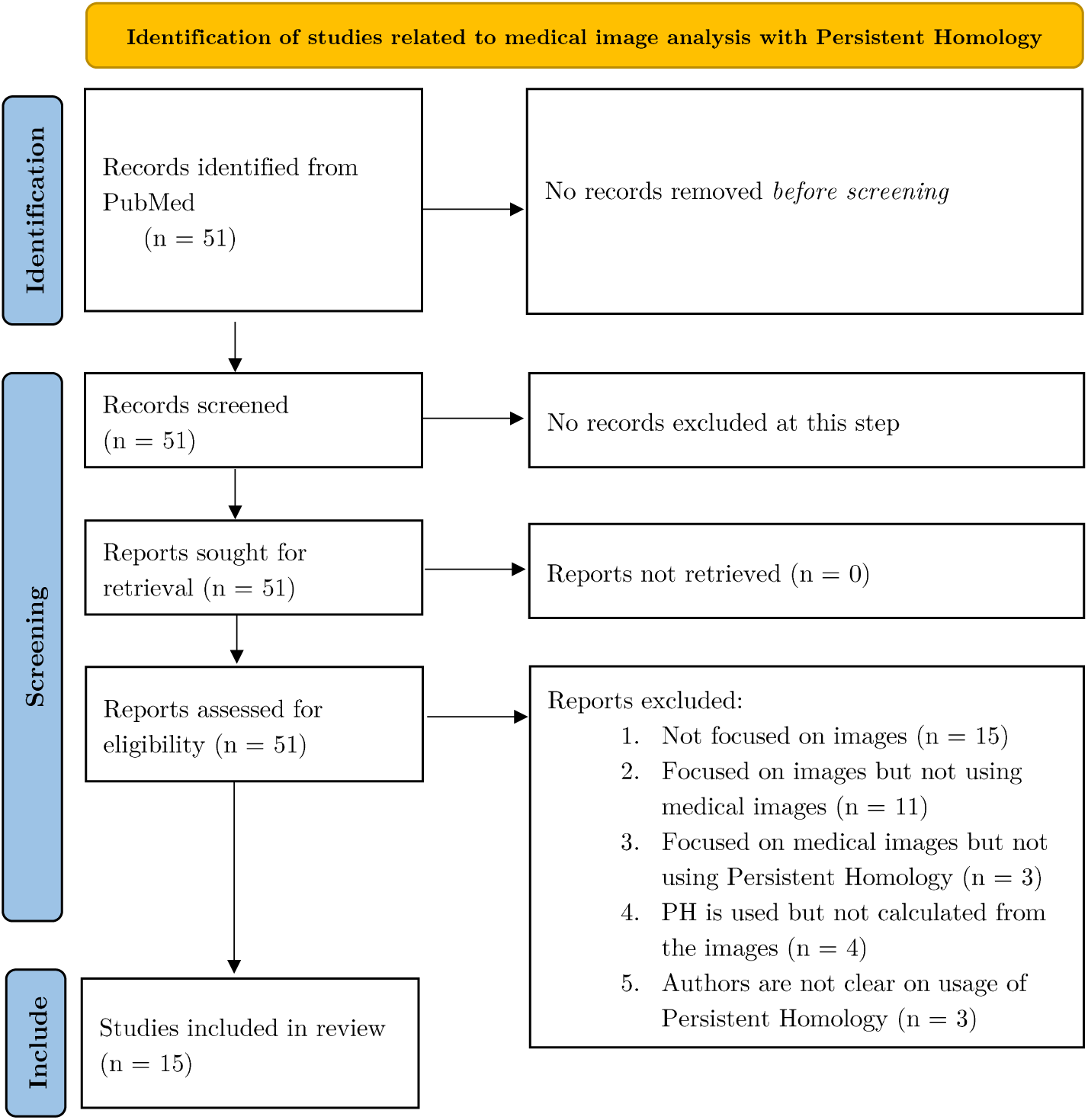
The PRISMA statement flowchart shows details of the steps taken to select studies which were included in the literature review.

The following research questions were formulated:

1. What filtration methodologies are applied to medical images to calculate Persistent Homology?
2. Can Persistent Homology improve performance of image processing algorithms?

Accordingly, the following search string was used:

“((Topological Data Analysis) OR (Computational Topology) OR (Persistent Homology)) AND ((microscop*) OR (medical imag*) OR (biomedical imag*)) NOT Review”

To exclude entries, the abstract was carefully analysed. Articles were excluded if the answer to any of the following questions was no:

- Does the given paper focus its research on images (as opposed to other data types, such as point clouds)?
- Are the images of a medical or biomedical nature?
- Does the paper use Persistent Homology as a key step in the image processing algorithm?

Then, the full text of the article was read. This closer inspection revealed that some papers were not considered to address the research questions. In addition, papers that lacked details about how PH was used were discarded.

## 4 Results

The initial search result returned 51 entries. After the initial screening, 24 entries were excluded, and in the final selection step, another 12 entries were discarded according to the steps described in Section 3. This process returned 15 records to be included in the systematic review (Table 1).

**Table 1.**
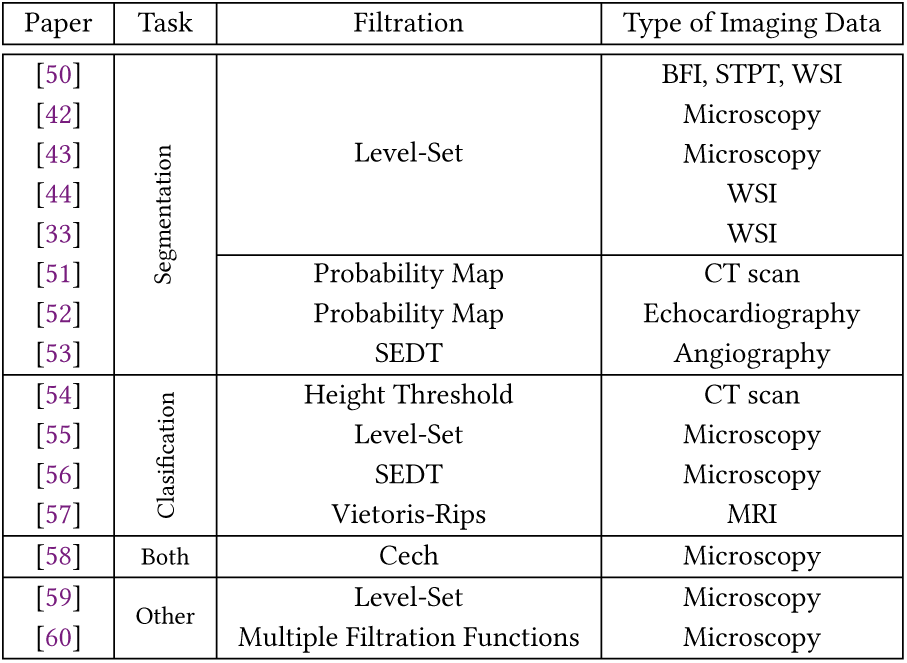
Full list of articles categorised by image processing task, filtration, acquisition technique: bright-field images (BFI), serial two-photon tomography (STPT), Whole slide imaging (WSI), Computed Tomography (CT), Magnetic resonance imaging (MRI).

Two main types of tasks using PH were identified, namely, image classification and image segmentation. Segmentation was performed in 8 studies (60%), classification in 4 studies (33.3%), both segmentation and classification in 1 study and 2 other papers performed tasks that cannot be considered segmentation or classification.

### 4.1 Segmentation by Level-Set Thresholding

Segmentation by level-set thresholding as previously described was used in 5 papers. This is by far the most common and straightforward way to make a filtration from a greyscale image. In brief, this process sequentially creates a sequence of complexes that form a filtration by lowering (or raising) a threshold directly on the intensity of the pixels. In some cases, a filtration is used to train a CNN through a topological loss function [42, 51].

Level-set thresholding where *n* levels were used to create *n* binary images was used by Rojas-Moraleda [33]. The connected components of these images are then tracked as the threshold varies in order to calculate their persistence, this helps identify individual objects, like cells, which may be very close and appear as merged into one with traditional segmentation methods.

Haft-Javaherian *et al.* applied a threshold directly on 3D volumes to build a filtration based on the connected components of the resulting binary volumes [42]. Using the resulting filtration, a topological loss-function encoding the target Betti Numbers was combined with a customised cross-entropy function from the DeepVess neural network architecture to train a neural network [61]. Similarly, Shin *et al.* also combined PH with Deep CNNs to perform segmentation [51]. This was done by inputting CT scans of the Small Bowel to a CNN that outputted a 2D probability map, which was then sequentially thresholded to build a filtration and obtain a Persistence Diagram. The births and deaths of the points in the diagram were used to define a loss function that was used to fine-tune a pre-trained U-net. Qaiser *et al.* introduced a novel concept known as Persistent Homology Profiles (PHPs) used as features built by level-set thresholding patches of greyscale 2D images [44]. The PHPs encode the 0 and 1-dimensional Betti Numbers at each threshold, as well as their ratio, as a real-valued function of the threshold. These features were then used to train a deep Convolutional Neural Network (CNN) to perform classification of tumour vs non-tumour patches. The final segmentation was given by the pixels in those patches that were classified as being part of a tumour. This description forms the base for the two algorithms built by Qaiser *et al.*, the first is known as the *Fast* algorithm while the *Accurate* algorithm added deep convolutional features into the feature extraction and segmentation process.

Banerjee *et al.* combined PH with Discrete Morse Theory (DMT) [62] [63] to perform segmentation on Bright-Field (BFI), Fluorescent Whole Slide (WSI), and Fluorescent Serial Two-Photon Tomography (STPT) images of the brain with tracers [50]. Initially, images were smoothed with a Gaussian filter. Then, a level-set filtration was built from the smoothed images. DMT was used to track the formation and merging of components and holes, which were used to calculate persistence. Less persistent features were discarded and the final segmentation was obtained by combining the topological features with a Deep CNN architecture known as ALBU [64].

Panconi *et al.* employed a human-in-the-loop approach for their segmentation model [43]. First, a level-set filtration was obtained from greyscale images to produce an initial segmentation. Since objects of interest tend to be brighter than the background (in this case) and merge with more persistent areas later in the filtration than noise does, the segmentation was simply made by thresholding for connected components that have a high birth value and a high persistence. These thresholds were manually raised or lowered to match the expected number of connected components in the greyscale image. This initial segmentation was used to isolate inner and outer sections of cells and structures with a variation of the Swinging Arm method [65]. The algorithm was tested on synthetic data and microscopy images of HEK293 cells and the R265 strain of *C. gattii* cells.

### 4.2 Filtering without Level-Set Thresholding

Sun *et al.* segmented the myocardium in echocardiography images [52]. This article tackles two problems with existing CNN segmentation algorithms. First, a smooth boundary is sought, in a segmentation but this can lead to many inaccuracies. The second problem is that due to poor image quality, an automatically segmented myocardium can be broken up into two or more pieces, which is anatomically incorrect. Thus, the model used PH in two ways:

- PH was used to build a loss function that assigns weights given to certain pixels on the boundary of the myocardium to update the weights of a pre-existing segmentation model.
- PH was used to maintain the integrity of the myocardium. This means that the final segmentation must consist of a single connected component with no holes, while the background is expected to be one connected component with one hole.

In both cases, PH was calculated, separately, from the ground truth images and a 2D probability map. These 2D structures were first transformed into a 3D point cloud, where the *x*, *y* values of a point are given simply by its position on the image, while the height *z* of the point is given by its assigned pixel/probability value. On the point cloud, a filtration of Alpha Complexes is calculated [66]. This filtration does not require thresholding pixel intensities.

Molina-Abril and Frangi sought to improve segmentation of blood vessels through the use of PH [53]. Segmentations of blood vessels often yield an incorrect result with separate blood vessels that touch each other. The proposed method takes an initial segmentation, skeletonises it, and uses a measure known as the Kissingness Measure (KS) to identify critical points. This measure takes into account the average differences in radius and average angle formed between branches at intersection points. The centrelines with the higher KS form what the authors called the suspicious kissing set. To create a filtration, a spring force was applied to the initial segmentation towards the intersection point until there is a topological change around the point. The smaller the forces required to make this change, the higher the probability that the point will be selected as critical. An algorithm based on active contours is then used to remove the critical points [67].

In the article by Edwards *et al.* classification and segmentation were performed [58]. PH was used to classify between cells that have been treated with different perturbations that affect the actin cytoskeleton. The treatments used were small-molecule inhibitors and genetic modifications. Initially, small circular patches were taken from the original image and high-intensity pixels were selected inside the patches, along with evenly spaced pixels on the boundary of the patch. The pixels form a point cloud and a filtration of Cech complexes was created by varying a distance parameter [37]. Persistence landscapes were calculated from the filtration and used to train Support Vector Regression (SVR) classifiers [68]. The SVR classifiers classified patches individually. The classification of an image was given by the average label assigned to its patches by the SVR classifiers. The segmentation was created by mapping values assigned to patches by the SVR classifiers back to pixels on the image. When visualised on top of the original image, with negative-valued pixels as one colour and positive-valued pixels as another, the different colours overlap with areas where treatment has affected the cells.

### 4.3 Performance of segmentation procedures

Table 2 shows performance metrics reported by each author that dealt with a segmentation problem. All metrics were calculated at the pixel-level, unless stated otherwise (like the algorithms tested by [44]). It is important to note that in the case of [33], the authors did not explicitly state the *F*_1_ score, but it was possible to calculate through the confusion matrix provided. Additionally, [33] and [52] tested the corresponding methodologies on two different datasets and obtained one score for each, these scores were averaged to obtain a single *F*_1_ score for each algorithm.

**Table 2.** Segmentation models together with the performance metrics reported by the authors. ^(∗)^ : averaged from testing on two different datasets. ^(∗∗)^ calculated from confusion matrix. ^(∗∗∗)^ : tested on synthetic data.

| Model | Evaluation Metric |
| --- | --- |
| [50] | $F_1$: 0.86 |
| [51] | $F_1$: 0.85 |
| [42] | $F_1$: 0.87 |
| [52] | $F_1$: 0.82 (*) |
| [33] | $F_1$: 0.91 (*)(**) |
| [43] | Sensitivity: 0.88, Specificity: 0.88 (***) |
| [44] Fast | Patchwise $F_1$: 0.83 |
| [44] Accurate | Patchwise $F_1$: 0.84 |

From Table 2 it is seen that all algorithms except [52] reached an *F*_1_ score of 85 or higher, which indicates a sufficiently good segmentation. In the case of [50], the segmentation was compared against other state-of-the-art methods (U-Net [69], SegNet [70], ALBU [64]), outperforming all of them. As with the previous article, the authors in [42] compared their method against state-of-the-art CNN segmentation methods ([61, 69, 71–73]) outperforming all in Dice and Jaccard index. The method in [51] was compared with a decoder network without any topological information incorporated into the training process: it was found that the loss function with topological prior improves the segmentation performance. The method in [33] was not directly compared to other segmentation models, however, the obtained *F*_1_ score was above 0.9, which indicates an extremely accurate segmentation. The model presented in [52] was the only that did not reach a 0.85 in the reported pixelwise *F*_1_ score. However, it outperformed the state-of-the-art methods it was compared to (clDice [74], TopoNet [75], and U-Net [69]).

For [43] specificity and sensitivity were reported. From these metrics, it is impossible to calculate the *F*_1_ score or the Jaccard index without knowing the proportion of pixels of the object to the pixels of the background.

In [44], two different models were tested. In their methodology, a segmentation was produced by taking small patches from the complete image and classifying each patch into one of two classes. In this case, patch-level *F*_1_ was reported.

It is important to note that in [58] and [53] no segmentation performance metrics were reported. In [53] PH is used to detect errors in a pre-existing volumetric segmentation, therefore detection and correction rates are provided: 81.80% and 68.77%, respectively.

### 4.4 Classification

Unlike the articles where segmentation is performed, the classification models showed a much more diverse list of ways of making a filtration to calculate PH. Because of this, the models have not been grouped by filtration in this section. Belchi *et al.* studied 3D CT scans of patients’ lungs with the objective to characterise lung scans of patients with Chronic obstructive pulmonary disease (COPD) versus healthy patients, and smokers versus non-smokers [54]. A centreline was fitted to the 3D scans and metrics from this line were extracted via PH. A filtration was made by sequentially thresholding the centreline at different heights, creating a different graph at each height. The number of times the centreline changes vertical direction and the number of branching points were tracked. These metrics together with the length of the bronchial tree were enough to distinguish Healthy Non-Smokers from both COPD groups; and healthy smokers from patients with mild COPD through Kolmogorov-Smirnov tests.

Pritchard *et al.* used PH to study the internal microstructure of bone samples in [56]. The authors wished to identify differences in male and female groups of mice both with and without osteocalcin-expressing cells in TPaF and SHG images. First, greyscale images were binarised using Otsu’s method, and patches of the binary image were processed individually in the next steps [76]. A function called Signed Euclidean Distance Transform (SEDT) was applied to each pixel; it assigns the distance to the region of the opposite colour to each pixel, with black pixels assigned a negative sign. The transformed patch was thresholded sequentially to create a sequence of binary images. Similar to the level-set filtration, a vertex is added when a new region appears in the sequence of images and an edge is added between vertices if the corresponding pixels are neighbours (not including diagonals). The resulting persistence diagram was used to calculate statistical features. For each persistent statistic, hypothesis tests were done to estimate if two distinct groups give statistically different results. It was found that the statistics differ significantly for most permutations of the groups. Finally, a support vector machine (SVM) was trained on the statistical features to classify a brand-new patch [77].

Rammal *et al.* employ a typical level-set filtration [55]. The authors’ goal was to estimate the Gleason Score from Spatial Inference Light Microscopy (SLIM) images of cancerous prostate glands. Overlapping patches of the greyscale images were processed individually to obtain the following eight features in each: mean pixel intensity, standard deviation of pixel intensity, mean lifespan of components and holes, standard deviation of the lifespan of components and holes, the persistent entropy for components and holes. The latter six are all topological features. Details on persistent entropy can be found in [78]. The features were used to train five different ML methods for 5-class classification (one class per possible Gleason Score). A decision tree classifier was the highest performer.

Xin *et al.* use PH *recursively* to diagnose and provide prognosis of Multiple Sclerosis (MS) through lesions appearing in MRI scans of the human brain [57]. Lesions visible on brain MRI scans were converted to a point cloud in 3D space by matching the centroid of each region to a point. A filtration was obtained via the Vietoris-Rips filtration [37], this filtration was called the G-Net. Taking the G-Net, a Community-Level network (C-Net) was obtained by matching *k*-simplices in the G-Net to vertices in a graph *G*. Edges between vertices in *G* are drawn if the corresponding *k*-simplices in G-Net share a *k* − 1-simplex. This yields a simpler representation of the local topology of the lesions. A filtration in C-Net naturally arises as simplices appear and fuse in the G-Net - the lifespan of a feature is given by the steps in the filtration where the members of a community (the vertices) do not change. Using the C-Net, statistical features from each community were obtained and used to train two different classifiers to distinguish between MS and healthy patients.

As mentioned earlier, in the article by Edwards *et al.* classification was also performed [58]. The details can be found in 4.1.

### 4.5 Performance of classification procedures

In image classification tasks, reported metrics are more standardised than in segmentation. Even though some authors may report other metrics as well, this section focuses solely on the ROC-AUC (Receiver Operating Characteristic - Area Under the Curve) and *F*_1_ score of the predictions. Table 3 shows the results reported by the authors that dealt with an image classification task.

**Table 3.** Classification models and ROC-AUC, *F*_1_, metrics reported by the authors. ^(∗)^ : averaged from testing on twelve different datasets.

| Algorithm | ROC-AUC | $F_1$ |
| --- | --- | --- |
| [58] | 0.97<sup>(*)</sup> | 0.94<sup>(*)</sup> |
| [56] | - | 0.78 |
| [55] | 0.99 | - |
| [57] | 0.88 | 0.87 |

Notably, the scores reported for the model in [58] are very close to 1. This indicates an outstanding performance in classification. It should be noted that the results were averaged from scores obtained on twelve different datasets.

However, the lowest *F*_1_ obtained for an individual dataset was 0.816, on which it also reached a very high ROC-AUC of 0.908. This was not compared against other models.

The method in [56] used an SVM to test whether topological features are enough to distinguish between control and test populations. Two different datasets were used (from two different image acquisition techniques). An *F*_1_ score of 0.78 was obtained when averaged over both datasets.

An extremely high ROC-AUC of 0.99 was achieved in [55] when the features were classified using a Decision Tree Classifier (DTC). The highest accuracy obtained by the DTC was 95%.

The previously mentioned classification models were not compared with other state-of-the-art models on the same datasets. This contrasts with the model presented in [57], which was tested against other models based on PH. An ROC-AUC of 0.88 and an *F*_1_ score of 0.87 were obtained, which means that this model outperformed all the others it was compared to.

Notably, no performance metric was reported in [54]. However, Kolmogorov-Smirnov tests revealed that topological features were statistically significantly different between healthy smokers and moderate COPD patients, as well as between healthy non-smokers and moderate COPD patients.

### 4.6 Other types of Analyses

Another class of articles did not perform any segmentation or classification of images but instead used PH to aid in other tasks. In this subsection those articles are presented.

Koseki*et al.* studied skin function looking to make a regression model that predicts Transepidermal Water Loss (TEWL) [60]. Greyscale images of the skin were binarised using Otsu’s method, then downsampled. Three different filtration functions were tested: k-Nearest Neighbour (k-NN) function, signed distance transform, and level-set thresholding. The k-NN function assigns to each pixel a value based on the distance to the k-th nearest white pixel. The signed distance transform assigns to each white pixel, the Manhattan distance to the nearest black region and, conversely, to each black pixel, the negative Manhattan distance to the nearest white region. In each case, the values assigned to the pixels by the functions were thresholded sequentially, creating binarised images and a filtration from the sequence. PH features were stored as a vector known as a Persistence Image [79]. Combined with other features such as age, sex, temperature, and humidity, several ML methods were tested to predict TEWL.

Herron *et al.* aimed to find the structure and organization of phagocytic podosomes [59]. This was done by first culturing Macrophages and introducing Immunoglobulin G (IgG) to induce the formation of podosomes with well-defined geometry. PH was used to localise the podosome sites in images of the cultures. Images were acquired using Structured Illumintation Microscopy (SIM) and Interferometric Photoactivated Localization Microscopy (iPALM). To find the podosome sites, the image was first smoothed using a Gaussian filter and small Gaussian noise was also added so that each pixel became distinct from its neighbours. A level-set filtration was built and a persistence diagram was obtained from the filtration, then *k*-means clustering was used to distinguish between relevant features and noise. The steps described previously were done twice, the first application uses a small Gaussian kernel, and was used to calculate holes to locate podosomes; while the second application used a larger Gaussian kernel and was used to calculate 1-dimensional holes to locate phagocytic sites. After this, some extra refining steps were done to improve the analysis. Podosomes that were far away from a phagocytic site were ignored, phagocytic sites that were only associated to 2 podosomes or less were also ignored, then the centre-point of the site was determined by the barycentre of all the podosomes associated to it.

### 4.7 Performance of other types of analyses

Due to the differences in the tasks performed by the models that did not deal with Classification or Segmentation, standard metrics were not compared. In [54], regression was performed using topological features obtained from images as independent variables to predict TEWL. The best model obtained an *R*^2^ of 0.524 and a Root Mean-Squared Error of 3.0 was obtained, indicating a sufficiently good fit. In the case of the model for localization of podosomes in [59] was compared against manually identified phagocytic podosomes, obtaining a false discovery rate of 0.01.

## 5 Conclusion

In this review, an in-depth analysis of 15 different articles was performed. These articles all use PH in some key step of a model used to analyze medical images, for different tasks such as segmentation, classification, or other applications. It is worth highlighting that, in those papers where a performance metric was reported, the models obtained a sufficiently high score, independently of the task it was used for. In most cases, PH is used to extract multi-scale topological features from images that perhaps other algorithms are not able to. The high-scoring performance metrics obtained by the models underscore the fact that these topological features provide information which is beneficial to existing models used to segment or classify images.

Additionally, this review showcases the diversity of applications of PH. This is mostly shown by the two papers presented in subsection 4.6. While segmentation and classification are classic image analysis tasks, in [59] the structure and organization of podosomes is described using PH, while in [60], topological features obtained with PH are inputted into a linear regression model to predict TEWL from images. The first of these two papers showcases the power of topology to describe the overall shape of data. The second, demonstrates that PH can capture useful features from an image of skin that add information to regression models.

In subsections 4.1 and 4.4 it is seen that PH is applied mainly to image segmentation tasks, with sufficiently good performances. The main way that PH captures features from images to perform segmentation is through building a level-set filtration from the pixel intensities in images or patches of a whole image. Through thresholding, this filtration captures the most persistent objects in greyscale images while discarding regions that are essentially noise. In classification, the reported performance metrics are also high compared to other state-of-the-art methodologies. For this task, there is no prevalent way to build a filtration, such as the level-set filtration used in segmentation. Here, a larger variety of ways to build a filtration is found. In the case of [56], SEDT is first applied to previously binarised images, this function is then thresholded to build a filtration. Xin *et al.* in [57] filter the Vietoris-Rips complex to build a filtration [37]. Then, this same filtration is used to build a higher-level filtration that captures the dynamics of local clusters (communities) of simplices in the filtration.

It is worth mentioning that PH does present some downsides/challenges. For example, PH requires high-level understanding of topological concepts in order to build on existing ideas. However, this high-level understanding is not necessary to perform analysis with PH. Many tools and libraries are available to start using PH in programming languages and software such as: Ripser for Python [80], the Topology ToolKit (TTK) for Python and ParaView [81], SNT for ImageJ and Fiji [82]. Another challenge that PH presents is the complexity of constructing a filtration. For large datasets (point clouds or images), constructing a filtration to calculate PH can become a time-intensive task.

Considering the above, the benefits of integrating PH into image processing pipelines seem to outweigh the challenges. For this we present the following arguments:

1. As mentioned previously, PH can extract features from data that may not be initially attainable by other methods. These types of features can easily be used as priors to improve existing image processing pipelines.
2. The most popular methods for image processing recently have been those that involve CNNs. PH can be easily integrated into CNNs by adding a topological term to the loss function, as seen in [42] and [51].
3. The calculation of Betti numbers (and thus PH) becomes a relatively simple task once a filtration is built. This can easily be done by representing the simplicial complex as a Boundary Matrix. Betti numbers can be found by using simple matrix multiplication (which is very fast) [24].
4. In this review, it has been shown that PH can help improve the performance of image processing models. Whether it is compared with SegNet, U-Net, and ALBU, as shown by Banerjee *et al.* [50], or demonstrates clear improvements over a CNN without a topological prior, as shown by Shin *et al.* [51]. It is evident that PH provides benefits when integrated into image processing models.

### 5.1 Future Directions

Although significant progress has been made in the application of PH to medical image analysis, there are still gaps in the field. Based on the findings of this literature review, it is possible to draw investigation paths that still require further research:

- **Classifying histology patches:** the example in Section 2.4 can be expanded into a complete classification model. As seen in Figure 7, statistical attributes of the persistence diagrams show distinct distributions across samples of tissue types. These attributes are similar to the ones computed by Pritchard *et al.* [56]. These could be used to train an ML model for tissue classification.
- **Analysis of topological features in disease progression over time:** many diseases present with structural changes as time progresses. PH presents a way to track the topological changes over different time points. Potential applications could include quantifying gradual structural changes in brain MRIs due to neurodegenerative diseases such as Parkinson’s or Alzheimer’s, analysis of how tumors evolve over time in response to treatment, or tracking structural changes in heart tissue over time.
- **Application of PH in 3D and 4D medical imaging:** as shown in this review, most PH-based studies focus on 2D medical images, but many imaging modalities produce 3D volumes. PH can be extended to higher-dimensional datasets by considering higher-dimensional holes. However, the effectiveness of using PH features in these datasets is still under-researched. A direct application where this idea might be useful is in volumes of Electron Microscopy images [32]. Dark spots such as mitochondria in a volume of HeLa cells could potentially be captured by the level-set filtration as they resemble holes inside a cell.
- **Sensitivity to parameters:** when building a filtration, some parameters are arbitrarily chosen. For example, in the level-set filtration, usually all possible pixel intensity values are chosen as the level sets (*e.g.*, all values in the rage [0, 255]). However, reducing the number of levels (*e.g.*, six different pixel intensities as in Figure 4) could have an effect on the persistence diagrams and classification accuracy. Additionally, different filtration functions (*e.g.*, SEDT) transform the image in different ways, highlighting different parts and features of the images. Yet, to the best knowledge of the authors, no studies systematically compare their effects on the same classification or segmentation task. For example, an intensity-based filtration may highlight cell density variations, while distance-based filtrations (*e.g.*, SEDT) may emphasize spatial organization. Comparing these effects could refine topological biomarkers for cancer diagnosis. Additionally, in diseases like COPD, tissue degradation leads to structural changes in the bronchial tree, as shown in [54]. Combining these findings with descriptors derived from other filtrations could capture previously unknown aspects of lung texture and airways, helping in severity assessment.
- **Training classifying models using persistence image:** the Persistence Image (PI) encodes PH features into vectors [79]. Koseki *et al.* used PI for linear regression, but a similar approach coul be applied to classification and segmentation. For example, in the case of patches of histology WSI (Figure 6), the PI can be computed for each tissue type, which can be used to train a classification ML model (*e.g.*, Support Vector Machines, Logistic Regression, Deep Learning Classifiers).
- **Distance metrics between distributions:** as mentioned in the previous point, and seen in Figure 7, the statistical attributes of persistence diagrams vary according to the tissue classes. A simple way to compare the distributions is by the difference in their means. However, this approach does not take into account the standard deviations and overall shapes of the distributions. More complex distance functions such as the Bayes Discriminability Index [83], the Bhattacharyya Distance [84], Bhattacharyya Space [85] and the Kullback-Leibler Divergence [86] better capture the dissimilarities between distributions. Even more sophisticated measures, such as the Total Variation Distance [87] and the Wasserstein Metric [87], permit a rigorous mathematical quantification of differences between distributions. All these measures can be used to enhance statistical analysis and classification of tissue types by the scatterplots generated from the statistical attributes that each distribution yields.
- **Distance metrics between persistence diagrams:** distances between Persistence Diagrams can be mathematically defined in a similar way as the metrics between distributions. These distances include the Wasserstein Distance for Persistence Diagrams [88] and the Bottleneck Distance [41]. These metrics can be used to compare individual persistence diagrams but could also be extended to compare groups of persistence diagrams quantitavely. The results that these comparisons will have remain unknown but it is possible that these can help to quantify structural changes as an early biomarker to the progression of diseases such as cancer, or cardiovascular disease.

## Data Availability

All data produced in the present work are available upon request to the authors.

https://zenodo.org/records/1214456

